# Development and clinical evaluation of non-viral genome specific targeted CAR T cells in relapsed/refractory B-cell non-Hodgkin lymphoma

**DOI:** 10.1101/2020.09.22.20199786

**Authors:** Jiqin Zhang, Yongxian Hu, Jiaxuan Yang, Wei Li, Yue Tian, Guoqing Wei, Linjie Zhang, Kui Zhao, Yalei Qi, Binghe Tan, Mingming Zhang, Yi Li, Qiliang Tian, Chunqian Fang, Yuxuan Wu, Dali Li, Bing Du, Mingyao Liu, He Huang

## Abstract

In recent years, chimeric antigen receptor (CAR) T cell therapy has shown great promise in treating hematological malignancies. However, using virus in manufacture of CAR T cells brings about several problems. The application of CRISPR/Cas9 genome editing technology emerges in constructing novel CAR T cells by disrupting endogenous genes. Here we successfully develop a two-in-one approach to generate non-viral genome specific targeted CAR T cells through CRISPR/Cas9. By targeting a CAR in *AAVS1* safe harbor locus, we demonstrated that these CAR T cells behave comparable to those conventionally produced by lentivirus. Furthermore, *PD1*-knockin anti-CD19 CAR T cells show a superior ability to eradicate tumor cells with high PD-L1 expression. In the adoptive therapy for relapsed/refractory (r/r) aggressive B-cell non-Hodgkin lymphoma (B-NHL), we observed durable responses without serious adverse events and complete remission (CR) in patients treated with these *PD1* knockout CAR T cells. Collectively, our results prove the safety and feasibility of non-viral genome specific integrated CAR T cells, thus providing a new potential strategy for cancer treatment using these novel CAR T cells.

## Introduction

In these years, CAR T cell therapy has fast developed and showed great potential in cancer therapy, which is exemplified by the FDA approval of three anti-CD19 CAR T treatments^1-5^. Nevertheless, the involvement of virus in CAR T cell production causes several disadvantages. They include that the insertional mutagenesis increases the risk of tumor^6,7^. The specific responses to virus-derived DNA tend to impede CAR expression^8,9^ and the high expenses are required in virus manufacture^10^. Accordingly, some strategies, such as using transposon system^11-14^ and mRNA transduction^15,16^, are being exploited to generate CAR T cells without virus. A recent report showed that CRISPR/Cas9 technology could be applied in producing non-viral T cell products to correct point mutations or insert TCR element into a specific locus^17^. This prompts us to further optimize conditions to construct non-viral genome specific targeted CAR T cells. Here, we show that *AAVS1* locus specifically inserted CAR T cells perform as well as conventional CAR T cells. Moreover, *PD1* integrated anti-CD19 CAR T cells exhibit a robust ability to eliminate tumor cells in pre-clinical experiments. Importantly, our investigator-initiated clinical trials (IITs) further demonstrate the safety and effectiveness of these *PD1*-knockin CAR T cells in treating r/r B-NHL patients. Thus, we provide a new potent CAR T cell therapy for cancer treatment by using non-viral genome specific targeted CAR T cells.

## Results

First, we used fluorescent protein as target to optimize the protocol on producing non-viral genome specific integrated T cells (data not shown). Based on the optimal method, we chose *AAVS1* safe harbor^18,19^ for CAR targeting, which excludes the influence caused by functional endogenous genes, to evaluate whether this approach would affect the property of CAR T cells. Anti-CD19 CAR sequence was constructed, which was comprised of the intracellular domain of 4-1BB and CD3ζ(named as 19bbz). The integration efficiency of 19bbz in *AAVS1* is about 10% (up to 19.80%) on 7 days after electroporation (Figure 1a, b). The indel percentage ranged from 67%-87% in five representative donor cells by inference of CRISPR edits (ICE) analysis (Figure 1c). Also, the integration was unbiased among bulk CD3^+^, CD4^+^ and CD8^+^ T cells (Figure 1d). To understand the influence caused by the preparation per se, we comprehensively compared *AAVS1* integrated anti-CD19 CAR T cells (named as AAVS1-19bbz) with lentivirus-producing anti-CD19 CAR T cells (named as LV-19bbz). Although the electroporation procedure itself leaded to some cell damage, the final products had high cell viability after thorough recovery (Figure 1e). Interestingly, we found that electroporation manipulation conferred CD8^+^ T cells more growth advantage over CD4^+^ cells when compared to lentivirus infection (Figure 1f), which was consistent with the previous study^17^. It was observed that AAVS1-19bbz and LV-19bbz cells exhibited comparable cell expansion after tumor cell stimulation (Figure 1g, h). Our approach did not change the differentiation of T cell subsets (Figure 1i). In comparison to untreated T cells, AAVS1-19bbz cells well responded to tumor cells as LV-19bbz did, with a little differences on cell marker expression and cytokine secretion (Figure 2a, b). Importantly, like LV-19bbz cells, AAVS1-19bbz cells vigorously eradicated tumor cells in vitro and in vivo (Figure 2c-f). Taken together, these results demonstrated that the strategy to produce non-viral genome specific targeted CAR T cells is feasible.

**Figure 1.**
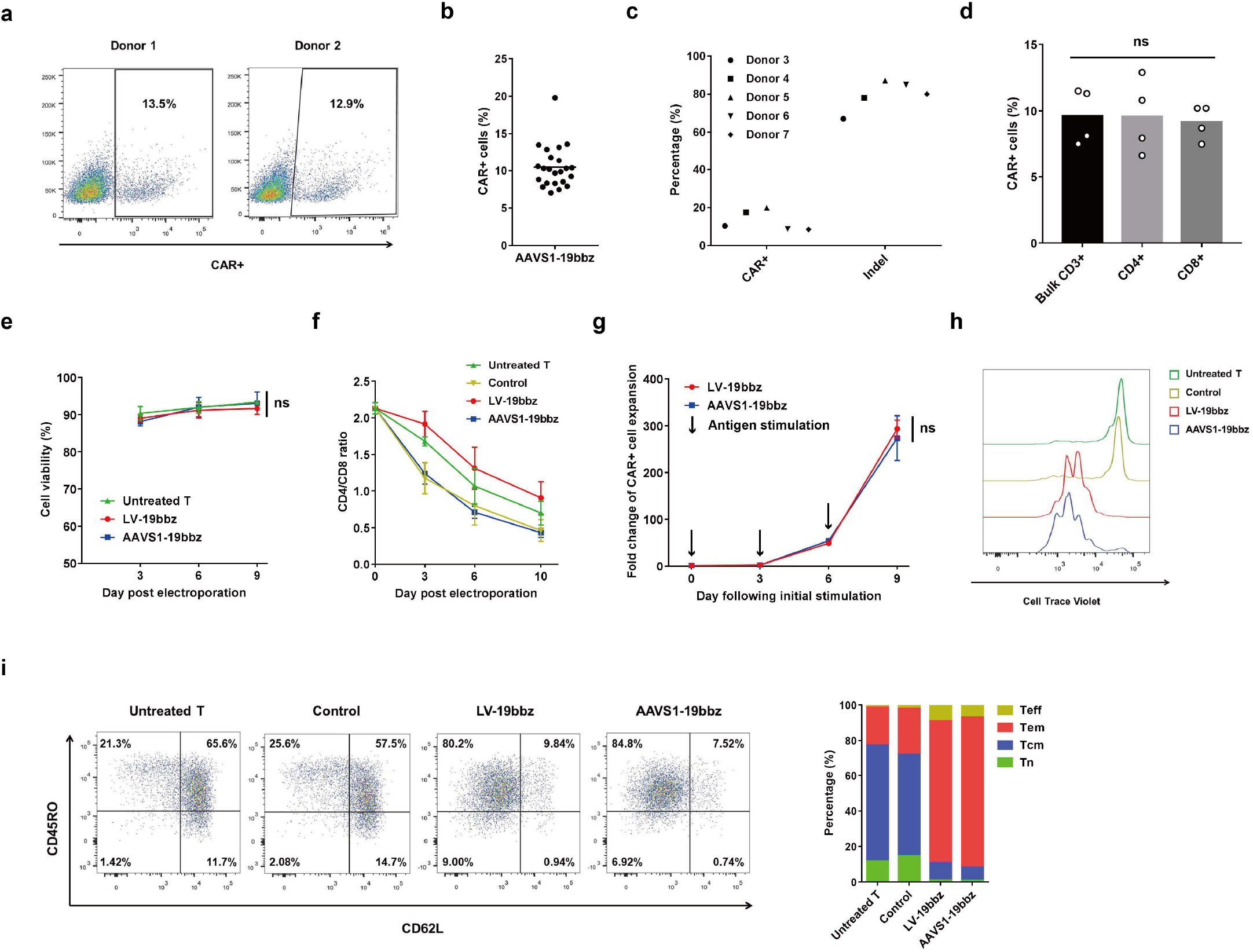
Non-viral *AAVS1* integrated CAR T cells behave comparable to conventional CAR T cells. a, CAR expression determined by flow cytometric staining 7 days after electroporation in two representative healthy donors. b, Percentage of CAR^+^ cells detected 7 days after electroporation (n=23 independent healthy donors). c, Percentage of CAR integration and *AAVS1* indel detected 7 days after electroporation in five representative healthy donors. d, Percentage of CAR integration in CD3^+^, CD4^+^ and CD8^+^ cells determined 7 days after electroporation (n=4 independent healthy donors). e, Cell viability detected by trypan blue staining on indicated days post electroporation. Data are mean ± SEM (n=3 independent healthy donors). f, Ratio of CD4^+^ and CD8^+^ cells on indicated days post electroporation. Data are mean ± SEM (n=3 independent healthy donors). g, Expansion of CAR^+^ cells after repeated stimulation with Raji cells. Data are mean ± SD (n=3 technical replicates). h, Representative histogram showing Cell Trace Violet staining of T cells after co-culture with mitomycin C-treated Raji cells for 5 days. i, Representative flow cytometry plots showing CD45RO/CD62L expression in T cells after 24 hour co-culture with Raji cells. The T cell subset differentiation is shown at right. Control samples were electroporated as AAVS1-19bbz cells except sgRNA addition. CD3^+^ (Untreated T, Control) or CD3^+^/CAR^+^ (LV-19bbz, AAVS1-19bbz) gated cells are analyzed in h, i. Mean value is shown in b, d. P values are calculated by one-way ANOVA (d) or two-way ANOVA (e, g).

**Figure 2.**
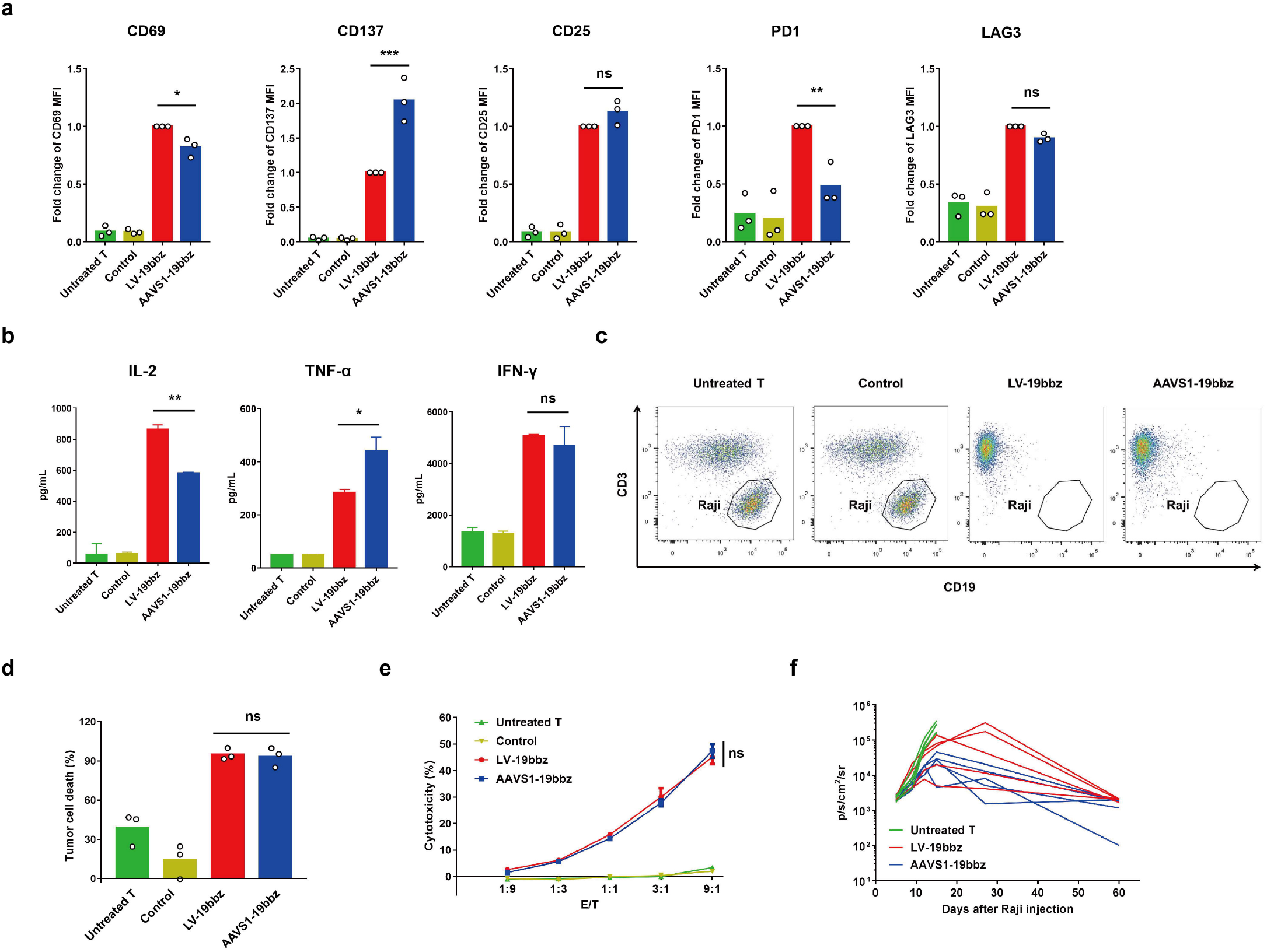
Non-viral *AAVS1* integrated CAR T cells effectively eliminate tumor cells as conventional CAR T cells. a, Median fluorescent intensity (MFI) of CD69, CD137, CD25, PD1 and LAG3 expression in T cells detected by flow cytometry after 24 hour co-culture with Raji cells (n=3 independent healthy donors). CD3^+^ (Untreated T, Control) or CD3^+^/CAR^+^ (LV-19bbz, AAVS1-19bbz) gated cells are analyzed. b, Representative result of cytokine secretion measured by bead-based immunoassay in the supernatant after co-culture with Raji cells for 24 hours. Data are mean ± SD (n=2 technical replicates). c, Representative flow cytometry plots showing lysis of Raji cells following 18 hour co-culture. d, The percentage of Raji tumor cell death detected by flow cytometry based cytotoxicity assay (n=3 independent healthy donors). e, In vitro cytotoxicity against Raji cells determined by LDH assay. E/T, effector/target. Data are mean ±SD (n=3 technical replicates). f, Bioluminescence kinetics of Raji tumor cell growth in NSG mice following different treatments (n=5). Control samples were electroporated as AAVS1-19bbz cells except sgRNA addition. Mean value is shown in a, d. P values are calculated by one-way ANOVA (a, b, d) or two-way ANOVA (e).

The blockage of PD1/PD-L1 pathway by inhibitors or gene editing has been reported to improve the antitumor activity of CAR T cells^20-23^. Accordingly, we set out to develop non-viral *PD1* targeted anti-CD19 CAR T cells (named as PD1-19bbz). The CAR expression of PD1-19bbz was detected in about 19% (up to 30.3%) donor T cells (Figure 3a, b). High indel percentage (83%-93%) was observed in total T cells from five representative donors (Figure 3c). The knockout of *PD1* in PD1-19bbz cells was demonstrated by low PD1 protein expression in CAR^+^ cells after co-culture with tumor cells (Figure 3d). It was noteworthy that PD1-19bbz cells expanded more than LV-19bbz cells after repeated stimulation by PD-L1 expressed Raji cells (Figure 3e). As indicated by other reports^24-26^, PD1 disruption did not affect the elevation of activation markers and cytokine secretion to counteract with target cells (Figure 3f, g). Notably, by contrast to LV-19bbz cells, PD1-19bbz cells showed more robust capability to clear PDL1-upregulated tumor cells in vitro and in vivo (Figure 3h, i). Collectively, we concluded that our non-viral PD1 integrated anti-CD19 CAR T cells function effectively.

**Figure 3.**
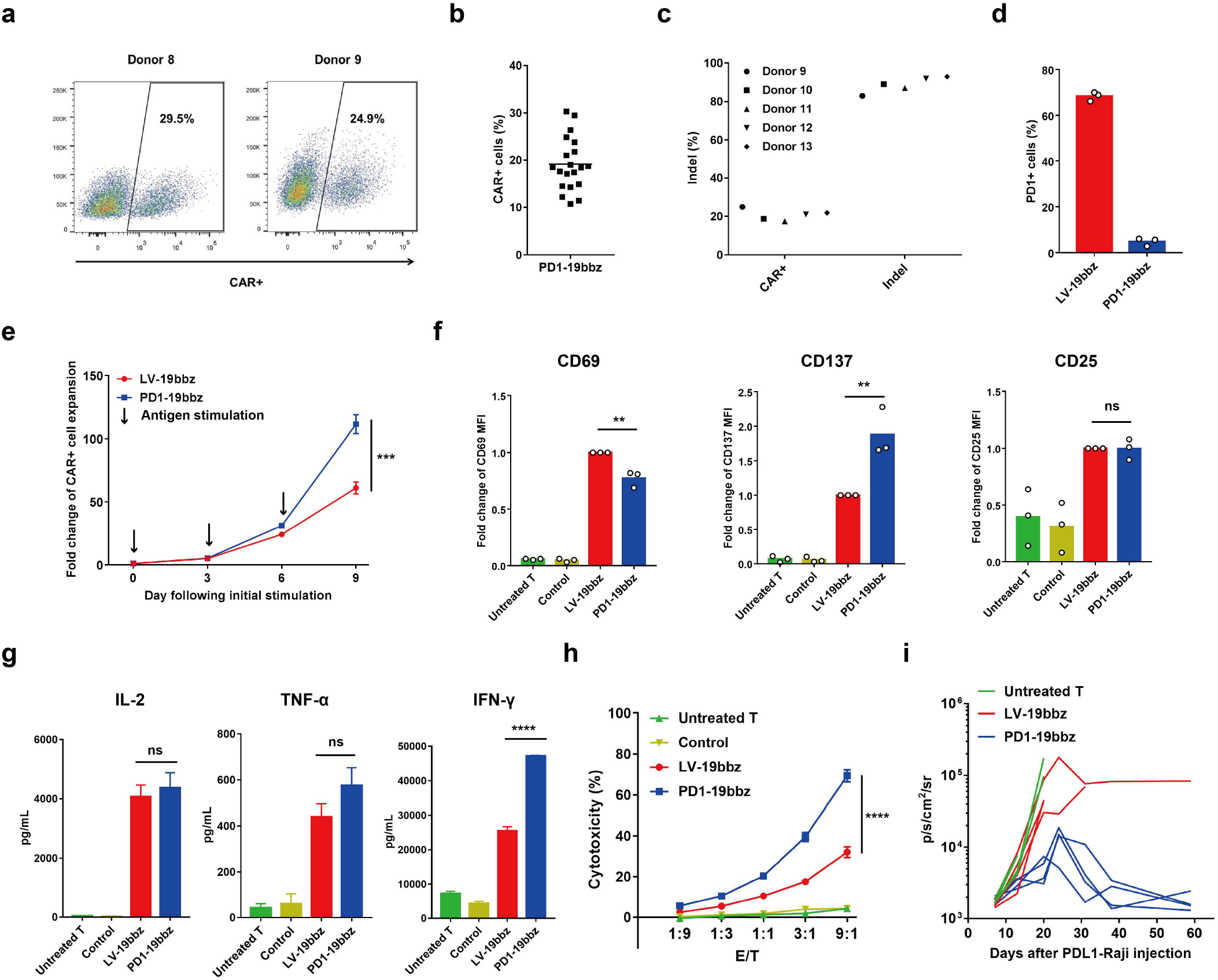
Non-viral *PD1* integrated CAR T cells outperform conventional CAR T cells. a, CAR expression determined by flow cytometric staining 7 days after electroporation in two representative healthy donors. b, Percentage of CAR^+^ cells detected 7 days after electroporation (n=20 independent healthy donors). c, Percentage of CAR integration and *PD1* indel detected 7 days after electroporation in five representative healthy donors. d, Percentage of PD1 expression detected by flow cytometry in CD3^+^/CAR^+^ gated cells after 24 hour co-culture with PD-L1 expressed Raji cells (n=3 independent healthy donors). e, Expansion of CAR^+^ cells after repeated stimulation with PD-L1 expressed Raji cells. Data are mean ± SD (n=3 technical replicates). f, MFI of CD69, CD137 and CD25 expression in T cells detected by flow cytometry after 24 hour co-culture with PD-L1 expressed Raji cells (n=3 independent healthy donors). CD3^+^ (Untreated T, Control) or CD3^+^/CAR^+^ (LV-19bbz, PD1-19bbz) gated cells are analyzed. g, Representative result of cytokine secretion measured by bead-based immunoassay in the supernatant after co-culture with PD-L1 expressed Raji cells for 24 hours. Data are mean ± SD (n=2 technical replicates). h, In vitro cytotoxicity against PD-L1 expressed Raji cells determined by LDH assay. Data are mean ± SD (n=3 technical replicates). i, Bioluminescence kinetics of PD-L1 expressed Raji tumor cell growth in NSG mice following different treatments (n=4). Control samples were electroporated as PD1-19bbz cells except sgRNA addition. Mean value is shown in b, d, f. P values are calculated by one-way ANOVA (f, g) or two-way ANOVA (e, h).

Based on our preclinical experimental data, we then proceeded to carry out IITs to evaluate the safety and feasibility of non-viral *PD1* integrated 19bbz cells in treating r/r B-NHL (ClinicalTrials.gov NCT04213469). Patient T cells were enriched from fresh peripheral blood mononuclear cells (PBMCs) via magnetic separation using anti-CD8/CD4 microbeads. After stimulated with CD3/CD28 agonist, PD1-19bbz cells were manufactured without virus by electroporation of recombinant spCas9 protein, a *PD1*-specific sgRNA and DNA template. The average percentage of CAR integration was about 20% and the highest percentage reached to 47.3%. The cell viability and indel percentage of final products exceeded 90% and 50%, respectively. All the patients were given a lymphodepleting chemotherapy using cyclophosphamide and fludarabine, followed by one infusion of PD1-19bbz cells with a dose of 0.5×10^6^-2×10^6^/kg body weight.

There were no CAR T cell related high-grade (⩾3) adverse events (AEs) after administration. Mild cytokine release syndrome (CRS) (grade 1) was observed and no neurologic toxicity occurred. In two representative patients, the expansion and persistence of CAR T cells showed similar change tendencies (Figure 4a, b). CR was achieved as shown by positron emission tomography–computed tomography (PET– CT) scans and it was ongoing at the time of last follow-up (90 days) (Figure 4c, d). Together, these data demonstrated the safety and feasibility of non-viral genome specific integrated CAR T cells in the clinical trials.

**Figure 4.**
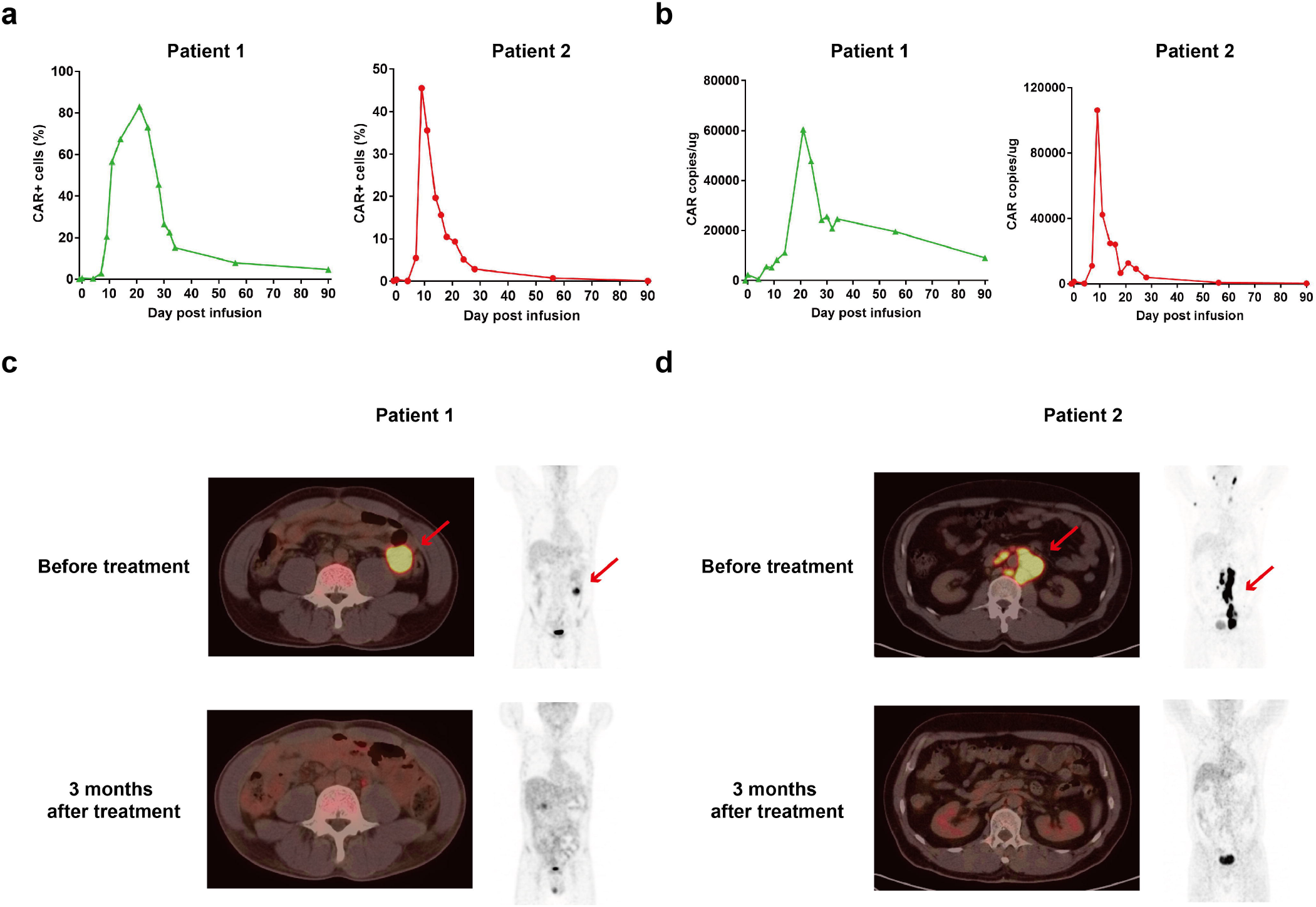
Non-viral *PD1* integrated CAR T cells potently eliminate tumor cells in patients with r/r B-NHL. a, Percentage of CAR^+^ cells in peripheral blood T cells of two representative r/r B-NHL patients on indicated days before and after infusion. b, CAR copy number in genomic DNA from peripheral blood of two representative r/r B-NHL patients is shown on indicated days before and after infusion. c, d, PET–CT scans for patient 1 (c) and patient 2 (d) before (baseline) and 3 months after CAR T cell infusion. Red arrows indicate the tumor lesions.

## Discussion

Here, we report a novel approach to generate genome specific targeted CAR T cells without virus. *AAVS1* inserted anti-CD19 CAR T cells well response to and effectively eradicate tumor target cells as those conventionally produced by lentivirus, demonstrating this strategy is effective. Furthermore, we developed a *PD1*-knockin anti-CD19 CAR T cell product and proved that it has superior potency to clear PD-L1 expressed tumor cells. It is of importance that manufactured PD1-19bbz cells show good safety and efficacy in treating patients with r/r B-NHL. Our approach combines both advantages of non-viral manufacture process and precise genome editing. Without using virus, it increases the safety and efficacy of CAR T cell product and markedly reduces the high cost. Locus-specific integration augments the homogeneity of CAR T cells and makes it possible to exploit versatile products.

Despite relatively low percentage of CAR^+^ cells in comparison to conventional CAR T cells, we substantiated that it does not influence the function of non-viral genome specific integrated CAR T cells and an additional enrichment step is unnecessary for clinical application. Given the insertion efficiency varies greatly between different positions, site screening is indispensible in advance. In addition, the length of target sequence for integration is another limitation. Further study aiming to raise the integration efficiency in T cells is needed to solve these disadvantages. On the other hand, although the electroporation step results in some cell damage, our data indicate that T cell expansion ability is not impaired and the cell number and viability of final product can fully meet the requirement of clinical administration. We find that while AAVS1-19bbz and LV-19bbz cells have comparable capability of killing tumor cells in preclinical assays, they indeed differ in gene expression and cytokine release when responding to antigen, which may be attributed to the difference of production process. It is also meaningful for performing clinical trials to evaluate non-viral *AAVS1* integrated CAR T cells in the future.

In accordance with the results of preclinical experiments, *PD1* integrated anti-CD19 CAR T therapy appears to show remarkable efficacy in treating r/r B-NHL in the clinic. Further analysis can be taken to validate whether *PD1* knockout indeed improves the potency of CAR T cells. It is also valuable to carry out more clinical trials with low-dose administration to comprehensively assess the efficacy of PD1-19bbz cells in the future. Consistent with two recently reported clinical trials^27,28^, we have not observed serious adverse events, especially high-grade CRS, during CAR T treatment, thereby further demonstrating the safety of CRISPR/Cas9 application in T cell therapy.

Taken together, we describe an innovative technology to develop non-viral genome specific targeted CAR T cells by CRISPR/Cas9 and demonstrate the safety and feasibility in preclinical experiments and clinical trials. Therefore, we pave the way for new application of gene editing in CAR T cells and provide a potent CAR T therapy, especially for immunosuppressive tumor.

## Data Availability

All data that supported the findings of this study are available from the corresponding author upon reasonable request.

## Methods

### Clinical trial information

This clinical trial was registered with ClinicalTrials.gov (NCT04213469). All patients provided written informed consent in accordance with the Declaration of Helsinki before enrolling in the study. The clinical protocol was reviewed and approved by the Clinical Research Ethics Committee of the First Affiliated Hospital, College of Medicine, Zhejiang University. CAR T cells for clinical therapy were manufactured by Bioray Laboratories Inc. Characteristics and clinical responses of two representative patients are shown in Table 1.

**Table 1.** Representative patient characteristics and clinical responses

| Patient number | Sex | Age | Diagnosis | Prior therapy | CAR<sup>+</sup> T cell dose (per kg body weight) | Best response | CRS | Neurological toxicity |
| --- | --- | --- | --- | --- | --- | --- | --- | --- |
| 1 | Male | 44 | DLBCL 2018 | R-CHOP<br>Sintilimab + ICE<br>R-Gemox + Chidamide<br>radiation therapy<br>Rituximab + Lenalidomide<br>Chidamide<br>total gastrectomy | 0.56×10<sup>6</sup> | CR | Grade 1 | None |
| 2 | Male | 53 | DLBCL transformed from LPL 2018 | R-COP<br>RCD<br>Lenalidomide<br>R-DAEPOCH | 2.04×10<sup>6</sup> | CR | Grade 1 | None |

DLBCL, Diffuse large B-cell lymphoma; LPL, Lymphoplasmacytic lymphoma; R-CHOP: rituximab, cyclophosphamide, doxorubicin, vincristine, prednisone; ICE: ifosfamide, carboplatin, etoposide; R-GemOx: rituximab, gemcitabine, oxaliplatin; R-COP: rituximab, cyclophosphamide, vincristine, prednisone; RCD: rituximab, cyclophosphamide, hexadecadrol; R-DAEPOCH: rituximab, etoposide, vincristine, doxorubicin; CR: complete remission

### Cell lines

Raji cells were purchased from ATCC and maintained in RPMI1640 medium (ThermoFisher) supplemented with 10% fetal bovine serum (ThermoFisher). Firefly luciferase (ffLuc) stably expressed Raji cell line was established by lentivirus infection. PD-L1 stably expressed Raji cells were generated using lentivirus vector containing a co-expression cassette for PD-L1 and ffLuc. All the stable cell lines were underwent puromycine selection.

### Isolation and expansion of human primary T cells

Fresh PBMCs from healthy donors were purchased from Shanghai SAILY Biological Technology Co., Ltd. Fresh PBMCs from patients were collected by apheresis. PBMC was isolated by density gradient centrifugation using Ficoll (Sigma-Aldrich). T cells were enriched via magnetic separation using anti-CD8/CD4 microbeads (Miltenyi Biotech) and activated with T Cell TransAct (Miltenyi Biotech). T cells were cultured in X-VIVO media (Lonza) supplemented with 2% human AB serum or CTS™ Immune Cell Serum Replacement (ThermoFisher) and recombinant human IL-2 (100 units/mL), IL-7 (5 ng/mL) and IL-15 (5 ng/mL). Cells were harvested once the number reached the requirement of administration, and then washed, formulated and cryopreserved.

### CAR T cell generation by lentivirus

Anti-CD19 CAR cassette was composed of humanized single-chain variable fragment (scFv) derived from clone FMC63, the extracellular domain and transmembrane regions of CD8α, the intracellular domain of 4-1BB (CD137), and the intracellular domain of CD3ζ. The CAR sequence was cloned into the pCDH lentiviral vector backbone containing an EF1α promoter. Lentiviruses were produced by transfecting 293T cells with CAR plasmid, pMD2.G and psPAX2 using polyethylenimine (PEI). Virus supernatants were harvested after 3 days to infect primary human T cells.

### RNP production

One two-component single guide RNA (sgRNA) targeting *PD1* was chemically synthesized (GenScript) and resuspended with TE buffer. Ribonucleoproteins (RNPs) were produced by complexing *PD1* sgRNA and recombinant spCas9 (ThermoFisher) for 10 minutes at room temperature. RNPs were electroporated immediately after complexing.

### Human primary T cell electroporation

Electroporation was performed 2-3 days after T cell stimulation. The procedure was conducted following the manufacturer’s instructions by using Lonza 4D electroporation system. Briefly, pre-washed T cells were resuspended in the electroporation buffer P3. Meanwhile, RNPs were prepared followed by mixture with DNA template. Cells in electroporation buffer were then added and moved into electroporation cuvettes. Pulse code EO115 was chosen for electroporation. After electroporation, pre-warmed media was immediately supplemented and cells were transferred away from electroporation cuvettes.

### Flow cytometry

CAR and membrane protein expression was determined by flow cytometry. Cells were pre-washed and incubated with antibodies for 30 minutes on ice. After twice wash, samples were run on an LSRFortessa (BD Biosciences) and analyzed with FlowJo software. The following antibodies were used: FITC anti-human CD3, APC anti-human CD69, APC anti-human CD137, APC anti-human CD25, APC anti-human PD1, APC anti-human LAG3, BV421 anti-human CD45RO, APC anti-human CD62L, APC anti-human CD3, FITC anti-human CD19, FITC anti-human CD4, APC anti-human CD4, APC anti-human CD8 (All from BioLegend), PerCP-Cy™5.5 anti-human CD45 (BD Biosciences). For detection of CAR expression, biotinylated human CD19 (aa 20-291) protein (ACRO Biosystems) and PE Streptavidin (BioLegend) were added in order, or PE-labeled human CD19 (aa 20-291) protein (ACRO Biosystems) was used. For some experiments, CAR T cells were co-cultured with target cells at E/T 1:1 (AAVS1-19bbz involved experiments) or 1:2 (PD1-19bbz involved experiments) ratio for 24 hours before harvest. For detection of clinical samples, peripheral blood cells were stained with antibodies, followed by addition of Lysing Buffer (BD Biosciences) before running. CAR percentage was analyzed in CD45^+^/CD3^+^ gated cells.

### Indel percentage analysis

Genomic DNA was obtained using a DNA extraction kit (ThermoFisher). The fragments containing indel sites were amplified by PCR using specific primers and purified by a gel extraction kit (Tiangen Biotech). DNA sequencing was carried out and indel percentage was measured by ICE analysis (Synthego).

### CAR copy number analysis by qPCR

Blood samples were collected before and after CAR T cell infusion. Lysis Buffer (BD Biosciences) was first added and genomic DNA was acquired using a DNA extraction kit (ThermoFisher). A seven-point standard curve was generated by using 5×10^0^-5× 10^6^ copies/ul 19bbz lentiviral vector DNA. A pair of primers specifically targeting 19bbz sequence was used to measure CAR copy number in peripheral blood cells. qPCR was run on a QuantStudio™ 3 Real-Time PCR System (ThermoFisher). Each sample was determined in triplicate.

### Antigen stimulation and proliferation of CAR T cells

As antigen for stimulation, Raji or PD-L1 expressed Raji cells were pre-treated with mitomycin C (50 μg/ml) for 90 minutes at 37°C. CAR T cells were co-cultured with target cells at E/T 1:1 (AAVS1-19bbz involved experiments) or 1:2 (PD1-19bbz involved experiments) ratio for 3-4 days per stimulation. The number of CAR^+^ cells was enumerated by multiplying total cell number and CAR percentage detected by flow cytometry. Cell viability was measured by trypan blue staining.

### Cell Trace Violet proliferation assay

AAVS1-19bbz cells were labeled with Cell Trace Violet (ThermoFisher) according to the manufacturer’s instructions. Raji cells were pre-treated with mitomycin C (50 μg/ml) for 90 minutes at 37°C. CAR T cells and target cells were mixed at E/T 1:1 ratio. After 5 days, cells were harvested and run on an LSRFortessa (BD Biosciences).

### Bead-based immunoassay

CAR T cells were co-cultured with Raji or PD-L1 expressed Raji cells at E/T 1:1 (AAVS1-19bbz involved experiments) or 1:2 (PD1-19bbz involved experiments) ratio in the media without exogenous cytokines. The supernatant was collected after 24 hours and cytokines were measured using LEGENDplex™ bead-based immunoassay kit (BioLegend) according to the manufacturer’s instructions.

### Flow cytometry based cytotoxicity assay

AAVS1-19bbz cells were co-cultured with Raji cells at E/T 1:1 ratio for 18 hours. Flow cytometry was used to determine residual tumor cells by staining with APC anti-human CD3 and FITC anti-human CD19 antibodies. Cells were enumerated using CountBright™ Absolute Counting Beads (ThermoFisher) following the manufacturer’s instructions.

### LDH cytotoxicity assay

CAR T cells were co-cultured with Raji or PD-L1 expressed Raji cells at indicated E/T ratios. The cytotoxicity activity was measured by release of lactate dehydrogenase (LDH) using CytoTox 96® Non-Radioactive Cytotoxicity Assay (Promega) according to the manufacturer’s instructions.

### In vivo mouse experiments

All animal experiments conformed to the regulations drafted by the Association for Assessment and Accreditation of Laboratory Animal Care in Shanghai and were approved by the East China Normal University Center for Animal Research. For AAVS1-19bbz involved experiments, 6-to 8-week-old NSG male mice were injected intravenously with 2 × 10^5^ ffLuc-transduced Raji cells. CAR T cells were administrated intravenously at E/T 10:1 ratio after 5 days. For PD1-19bbz involved experiments, 6-to 8-week-old NSG male mice were inoculated intravenously with 5 ×10^5^ ffLuc-transduced PD-L1 expressed Raji cells. CAR T cells were injected intravenously at E/T 10:1 ratio after 10 days. Bioluminescence images were acquired and analyzed using IVIS Imaging System and software (PerkinElmer).

### Statistics

Experimental data are presented as mean ± SD or mean ± SEM as described in the figure legends. Data were analyzed by one-way ANOVA or two-way ANOVA as indicated using GraphPad software. A p value <0.05 was considered statistically significant. Asterisks used to indicate significance correspond to ***p < 0.001, **p < 0.01, *p < 0.05. NS, nonsignificance.

## Notes

### Competing Interest Statement

This work was paritally suppored by Bioray Laboratories Inc. Patents related to this manuscript have been applied.

### Clinical Trial

NCT04213469

### Funding Statement

This work was supported by National Key R&D Program of China (2019YFA0802802) and the National Natural Science Foundation of China (91857116, 31871453). This work was also paritally suppored by Bioray Laboratories Inc.

### Author Declarations

This research was reviewed and approved by the Clinical Research Ethics Committee of the First Affliated Hospital, College of Medicine, Zhejiang University reference number: 2020IIT(85)

## References

1 MacKay, M. et al. The therapeutic landscape for cells engineered with chimeric antigen receptors. Nat Biotechnol 38, 233–244 (2020).

2 June, C. H., O’Connor, R. S., Kawalekar, O. U., Ghassemi, S. & Milone, M. C. CAR T cell immunotherapy for human cancer. Science 359, 1361–1365 (2018).

3 June, C. H. & Sadelain, M. Chimeric Antigen Receptor Therapy. N Engl J Med 379, 64–73 (2018).

4 Tran, E., Longo, D. L. & Urba, W. J. A Milestone for CAR T Cells. New Engl J Med 377, 2593–2596 (2017).

5 Labanieh, L., Majzner, R. G. & Mackall, C. L. Programming CAR-T cells to kill cancer. Nat Biomed Eng 2, 377–391 (2018).

6 Michieletto, D., Lusic, M., Marenduzzo, D. & Orlandini, E. Physical principles of retroviral integration in the human genome. Nat Commun 10, 575 (2019).

7 Russo-Carbolante, E. M. D. et al. Integration pattern of HIV-1 based lentiviral vector carrying recombinant coagulation factor VIII in Sk-Hep and 293T cells. Biotechnol Lett 33, 23–31 (2011).

8 Atianand, M. K. & Fitzgerald, K. A. Molecular basis of DNA recognition in the immune system. J Immunol 190, 1911–1918 (2013).

9 Tao, J. L., Zhou, X. & Jiang, Z. F. cGAS-cGAMP-STING: The Three Musketeers of Cytosolic DNA Sensing and Signaling. Iubmb Life 68, 858–870 (2016).

10 Gandara, C., Affleck, V. & Stoll, E. A. Manufacture of Third-Generation Lentivirus for Preclinical Use, with Process Development Considerations for Translation to Good Manufacturing Practice. Hum Gene Ther Method 29, 1–15 (2018).

11 Kebriaei, P. et al. Phase I trials using Sleeping Beauty to generate CD19-specific CAR T cells. J Clin Invest 126, 3363–3376 (2016).

12 Monjezi, R. et al. Enhanced CAR T-cell engineering using non-viral Sleeping Beauty transposition from minicircle vectors. Leukemia 31, 186–194 (2017).

13 Hurton, L. V. et al. Tethered IL-15 augments antitumor activity and promotes a stem-cell memory subset in tumor-specific T cells. P Natl Acad Sci USA 113, E7788–E7797 (2016).

14 Maiti, S. N. et al. Sleeping Beauty System to Redirect T-cell Specificity for Human Applications. J Immunother 36, 112–123 (2013).

15 Lin, L. et al. Preclinical evaluation of CD8+anti-BCMA mRNA CAR T cells for treatment of multiple myeloma. Leukemia (2020).

16 Foster, J. B. et al. Purification of mRNA Encoding Chimeric Antigen Receptor Is Critical for Generation of a Robust T-Cell Response. Hum Gene Ther 30, 168–178 (2019).

17 Roth, T. L. et al. Reprogramming human T cell function and specificity with non-viral genome targeting. Nature 559, 405–409 (2018).

18 DeKelver, R. C. et al. Functional genomics, proteomics, and regulatory DNA analysis in isogenic settings using zinc finger nuclease-driven transgenesis into a safe harbor locus in the human genome. Genome Res 20, 1133–1142 (2010).

19 Oceguera-Yanez, F. et al. Engineering the AAVS1 locus for consistent and scalable transgene expression in human iPSCs and their differentiated derivatives. Methods 101, 43–55 (2016).

20 Cherkassky, L. et al. Human CAR T cells with cell-intrinsic PD-1 checkpoint blockade resist tumor-mediated inhibition. J Clin Invest 126, 3130–3144 (2016).

21 Rafiq, S. et al. Targeted delivery of a PD-1-blocking scFv by CAR-T cells enhances anti-tumor efficacy in vivo. Nature Biotechnology 36, 847–856 (2018).

22 John, L. B. et al. Anti-PD-1 Antibody Therapy Potently Enhances the Eradication of Established Tumors By Gene-Modified T Cells. Clin Cancer Res 19, 5636–5646 (2013).

23 Ren, J. T. et al. Multiplex Genome Editing to Generate Universal CAR T Cells Resistant to PD1 Inhibition. Clin Cancer Res 23, 2255–2266 (2017).

24 Rupp, L. J. et al. CRISPR/Cas9-mediated PD-1 disruption enhances anti-tumor efficacy of human chimeric antigen receptor T cells. Sci Rep-Uk 7 (2017).

25 Su, S. et al. CRISPR-Cas9 mediated efficient PD-1 disruption on human primary T cells from cancer patients. Sci Rep-Uk 6 (2016).

26 Guo, X. L. et al. Disruption of PD-1 Enhanced the Anti-tumor Activity of Chimeric Antigen Receptor T Cells Against Hepatocellular Carcinoma. Front Pharmacol 9 (2018).

27 Lu, Y. et al. Safety and feasibility of CRISPR-edited T cells in patients with refractory non-small-cell lung cancer. Nat Med 26 (2020).

28 Stadtmauer, E. A. et al. CRISPR-engineered T cells in patients with refractory cancer. Science 367 (2020).

